# Multiplex PCR assay for the Rapid detection of *Klebsiella pneumoniae* pathotypes

**DOI:** 10.1101/2025.05.08.25327209

**Authors:** Sanika Mahesh Kulkarni, Jobin John Jacob, T Praveen, V Aravind, R Subbulakshmi, S Preethi, Binesh Lal, Karthik Gunasekaran, Abi Manesh, Shraddha M Karve, J Sudarsana, Sanjay Bhattacharya, Anand Shah, Savitha Nagaraj, Priyadarshini Padaki, S Jayakumar, Renu Mathew, SM Rudresh, Shariqa Qureshi, S Nivedhana, Geethu Joe, Ekadashi Rajni, Kamini Walia, Balaji Veeraraghavan

## Abstract

**Introduction:** *Klebsiella pneumoniae* is a major cause of nosocomial infections, with its evolving pathotypes—including multidrug-resistant (MDR), hypervirulent (hvKp), and convergent strains—posing significant diagnostic and treatment challenges. The convergence of antimicrobial resistance and hypervirulence in clinical settings complicates timely identification, leading to difficult-to-treat infections with limited therapeutic options.

**Materials and Methods:** In this study, we designed and optimized a multiplex polymerase chain reaction (m-PCR) assay for the simultaneous detection of key biomarkers/genes associated with hypervirulent (*rmpA, rmpA2, iucA, peg344, iroB*), carbapenem-resistant (*bla-_NDM_, bla-_OXA-48-like_*), and convergent *K. pneumoniae* (hv-CRKp) pathotypes in clinical isolates. Specific primers were designed for these targets in a five-pair combination, with rigorous optimization of reaction conditions, including annealing temperatures, specificity, and sensitivity.

**Results and Conclusion:** The developed m-PCR assay exhibited 100% specificity when compared to whole genome sequencing data, successfully detecting all target genes without cross-amplification in ATCC control strains. The assay demonstrated high sensitivity, efficiently amplifying bacterial genomes from minimal DNA input as low as 1 ng/µL. Additionally, validation through sequencing confirmed the accuracy of detected amplicons.

This m-PCR assay offers a rapid, sensitive, and specific diagnostic tool for differentiating *K. pneumoniae* pathotypes in clinical settings, aiding in timely intervention and improved infection control measures.

**Impact Statement:** This study presents a multiplex PCR assay for the rapid and accurate detection of *Klebsiella pneumoniae* pathotypes, addressing the challenge of defining hypervirulence due to the diverse and evolving set of markers. By analyzing genomic data from public databases, the most well-established virulence and resistance markers were carefully selected to enable comprehensive strain differentiation. The assay simultaneously detects hypervirulent, carbapenem-resistant, and classical strains, offering a faster and more targeted alternative to conventional methods. Additionally, its adaptable design allows for modifications based on regional strain variations, making it a valuable tool for both clinical and epidemiological applications. This approach enhances infection control efforts and antimicrobial stewardship by enabling timely and precise pathogen identification, particularly in resource-limited settings. By bridging genomic insights with practical diagnostics, this assay provides a cost-effective, scalable solution to monitor and manage high-risk *K. pneumoniae* infections more effectively.

## 1. Introduction

*Klebsiella pneumoniae* (Kp), a Gram-negative, non-motile bacterium belonging to the Enterobacterales family, is known for its rapid dissemination, persistence, and phenotypic diversity (1,2). In hospital settings, Kp poses a significant threat, particularly to critically ill patients in intensive care units (ICUs), immunocompromised individuals, the elderly, and neonates (3,4). It is a frequent cause of severe infections, including pneumonia, urinary tract infections, sepsis, wound infections, and meningitis. Clinically, *K. pneumoniae* is classified into two primary pathotypes: Classical (cKp), which is responsible for nosocomial infections, and hypervirulent (hvKp), which is linked to community-acquired infections. cKp is further subdivided into multidrug-resistant (MDR-Kp) and carbapenem-resistant (CRKp) strains based on their antimicrobial resistance (AMR) profiles (5–8).

The prevalence of MDR *K. pneumoniae* in clinical settings has risen dramatically in recent decades. Studies indicate that approximately one-third of hospital-acquired Gram-negative infections are caused by Kp (9). In India, surveillance data from a multi-hospital network revealed that Kp accounts for 18% of bloodstream infections, with 57% of isolates exhibiting carbapenem resistance (10). Treating these infections is particularly challenging due to their resistance to most antibiotics, including last-line therapies. Consequently, the World Health Organization (WHO) has classified CRKp and extended-spectrum β-lactamase (ESBL)-producing strains as critical-priority pathogens (11). The frequent acquisition of virulence factors, metal resistance, and other horizontally transferred genetic elements further complicates the management of Kp infections (12,13).

Beyond nosocomial settings, hvKp has emerged as a significant community-acquired pathogen over the past two decades. Historically, hvKp strains exhibited a hypermucoviscous (hmv) phenotype, which was qualitatively assessed using the “string test” (14). This phenotype, along with enhanced virulence, stems from the overproduction of capsular polysaccharides and siderophores, driven by genes on the virulence plasmid (pLVPK), such as *rmpADC*, *rmpA2*, *iroBCDN*, and *iucABCDiutA* (15). In recent years, hvKp has evolved into multidrug-resistant hvKp (MDR-hvKp) by acquiring MDR plasmids. Conversely, MDR-Kp strains have integrated virulence plasmids, giving rise to convergent clones that exhibit both resistance and hypervirulence (16). These strains are most commonly reported in China, Southeast Asia, and East Asia, with increasing cases documented in Europe, the USA, and other Western countries (15–17). In India, a study from a tertiary care hospital identified 8% of isolates as belonging to this convergent pathotype (18). The emergence of CR-hvKp, harbouring resistance genes such as *bla*_KPC_, *bla*_NDM_, and *bla*_OXA-48-like_ has severely limited treatment options, often resulting in untreatable infections. The rising prevalence of MDR-hvKp in hospital settings is a pressing public health concern (19–22). Differentiating *K. pneumoniae* pathotypes is thus essential for assessing infection severity and devising effective treatment strategies.

Traditionally, hvKp identification relied on a positive string test (string >5 mm) and susceptibility to commonly used antibiotics. However, most convergent clones lack hmv and test negative in the string test, limiting its reliability as a specific marker for hvKp (23, 24). Genetic determinants of hypervirulence, such as aerobactin (*iucA*), metabolic transporter (*peg-344*), and regulator of mucoid phenotype (*rmpA*/*rmpA2*), have emerged as more robust biomarkers for PCR-based detection (25, 26) not all hvKp strains carry the same set of virulence genes, and frameshift mutations in *rmpA/rmpA2* can reduce the sensitivity of PCR assays (26–28). Given the urgent need for a rapid, comprehensive molecular tool to identify and monitor convergent strains carrying both resistance and virulence determinants, we evaluated multiple genotypic biomarkers to distinguish all three *K. pneumoniae* pathotypes: CRKp, hvKp, and CR-hvKp. Using these biomarkers, this study developed a multiplex PCR assay to accurately detect carbapenem-resistant hvKp and inform tailored treatment strategies for affected patients.

## 2. Material and Methodology

### 2.1 Genome sequence data retrieval

We have leveraged WGS data available at NCBI pathogen detection (https://www.ncbi.nlm.nih.gov/pathogens/) to identify carbapenem-resistant hvKp genomes. This database integrates genome sequences and annotates AMR and virulence genes present in bacterial genomes. Among the 52,502 assembled genomes of *K. pneumoniae* available in the database (dated 3^rd^ July 2023), we screened 12,846 genomes as hvKp by filtering for specific virulence genotypes (*rmpA/rmpA2* and*/*or *iucA*). Subsequently, genomes carrying carbapenemase genes such as *bla*_NDM_ and/or *bla*_OXA-48_ family, specific to Indian clinical contexts, were selected using various filter options within the database (https://www.ncbi.nlm.nih.gov/pathogens/refgene/). MLST, virulence genes, resistance, and virulence scores of genome assemblies were examined using Kleborate v2.4.1 (https://github.com/klebgenomics/Kleborate/releases/tag/v2.4.1) and BLASTn against reference genes downloaded from Institute Pasteur database (https://bigsdb.pasteur.fr/cgi-bin/bigsdb/bigsdb.pl?db=pubmlst_klebsiella_seqdef&page=downloadAlleles).

### 2.2 Primer design

For screening carbapenem-resistant hvKp, the coding regions of genes associated with hypervirulence (*rmpA, rmpA2, iucA*) were extracted from the genome assemblies (n=8177) through BLASTn searches locally, employing a sequence identity threshold of >80% and a coverage threshold of >80%. All reported *bla*_NDM_ and *bla*_oxa-48-like_ variant sequences in the NCBI reference gene catalogue (https://www.ncbi.nlm.nih.gov/pathogens/refgene/) were downloaded. Subsequently, the resulting FASTA files were aligned to reference sequences using the MAFFT program (https://github.com/GSLBiotech/mafft). Multiplex polymerase chain reaction (m-PCR) primers were designed for the primary assay, targeting conserved regions of *rmpA, rmpA2, iucA*, *bla*_NDM_, and *bla*_OXA-48-like_. Additional primers were developed for *peg-344* and *iroB*, allowing for flexibility in panel customization. Primer design was performed using the Thermo Fisher OligoPerfect Primer Designer (https://apps.thermofisher.com/apps/oligoperfect/). Selection criteria included a GC content close to 50%, a ΔTm of < 2°C, and a length of 18 – 22 bases. Before empirical testing, these primers were initially evaluated in silico against all target and non-target sequences using NCBI primer-BLAST (https://www.ncbi.nlm.nih.gov/tools/primer-blast/) and *in silico* PCR (https://insilico.ehu.es/PCR/). Specific primers and assays that target *bla*_KPC_ were adapted from previously published data (29). All oligonucleotide primer sequences used in this study are listed in Table 1 and were synthesized by Integrated DNA Technologies (IDT) Pvt Ltd, Singapore. Reference standard strains, as well as clinical isolates, were used for primer optimization and in vitro testing.

**Table 1:**
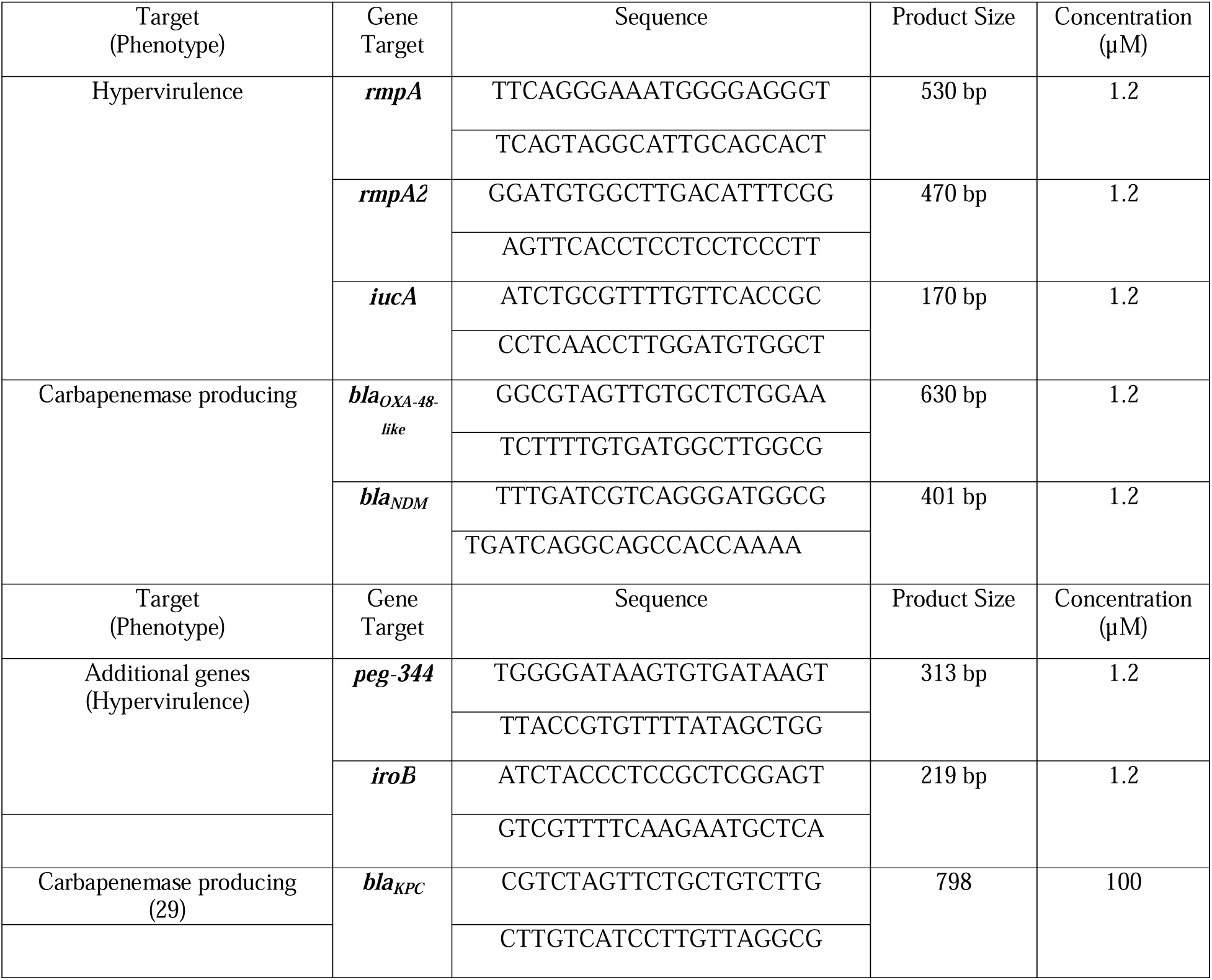
Oligonucleotide primers used in this study.

### 2.3 Characterization of bacterial isolates

*K. pneumoniae* strains (*n=150*) used in this study were clinical isolates collected between 2022 and 2023 at the Department of Clinical Microbiology, Christian Medical College, Vellore (n = 47), and from multiple centres across India, including Baby Memorial Hospital, Kozhikode (n = 76), Zydus Hospital, Ahmedabad (n = 10), Saveetha Medical College, Chennai (n = 5), St. John’s Medical College, Bengaluru (n = 5), BCMCH, Thiruvalla (n = 3), and TATA Medical Center, Kolkata (n = 4)(***Table S1***). These isolates were sub-cultured on MacConkey agar at recommended culture conditions. Isolates were identified and confirmed as *K. pneumoniae* by biochemical tests and MALDI TOF MS (VITEK® MS, bioMérieux). Screening for the hmv phenotype was conducted via the string test, following the described semi-quantitative method (1). A positive string test was defined by the formation of a mucoid string exceeding 5 mm in length upon contact with an inoculation loop.

### 2.4 Antimicrobial susceptibility testing

Antimicrobial susceptibility testing (AST) was performed by Kirby Bauer disc diffusion method according to CLSI 2022 and 2023 guidelines (30,31). The antimicrobial agents tested included Meropenem (10µg) and Ertapenem (10µg). Controls utilized for the testing included *Escherichia coli* ATCC 25922, *Enterococcus faecium* ATCC 29212, and *Pseudomonas aeruginosa* ATCC 27853.

### 2.5 DNA extraction and Whole genome sequencing

The study isolates (*n=150*) were cultured in LB broth (Oxoid, Hampshire, United Kingdom) at 37°C. Total genomic DNA was extracted from pelleted cells using the Wizard DNA purification kit (Promega, WI, USA). The concentration of extracted DNA was determined using NanoDrop One spectrophotometry (Thermo Fisher Scientific, MA, USA) and Qubit 3.0 fluorometry (Life Technologies, CA, USA), and the samples were stored at –20°C until further analysis.

A sequencing library was prepared using the Nextra DNA Flex library preparation kit (Illumina, San Diego, CA). Initially, genomic DNA (1µg) was fragmented, and adapters were ligated to the ends. The ligated DNA libraries underwent size selection, and the resulting products were PCR amplified with index primers following the manufacturer’s instructions. PCR amplified libraries were analyzed using the TapeStation 4200 (Agilent Technologies) with the High Sensitivity (HS) D5000 Screen Tape assay kit. Subsequently, the paired-end library was sequenced on a NovaSeq 6000 platform (Illumina, USA) at Unipath Specialty Laboratory Limited, Ahmedabad, India, generating 2 x 150-bp reads. Sequencing reads with a PHRED quality score below 20 was discarded, and adapters were trimmed using cutadapt v1.8.1. Quality assessment was performed using MultiQC (https://github.com/ewels/MultiQC).

### 2.6 Comparative genome analysis

The resulting high-quality reads were then subjected to assembly using Unicycler (https://github.com/rrwick/Unicycler). MLST, virulence genes, resistance, and virulence scores were examined using Kleborate (https://github.com/klebgenomics/Kleborate/releases/tag/v2.4.1). The assembled Klebsiella genomes were typed using the Ridom SeqSphere+ platform based on the core genome Multi Locus Sequence Typing (cgMLST) scheme for Klebsiella pneumoniae/variicola/quasipneumoniae available at (https://www.cgmlst.org/ncs/schema/Kpneumoniae2267/). A total of 2358 loci were selected and alleles were called using the chewBBACA algorithm (https://github.com/B-UMMI/chewBBACA). Minimum spanning trees (MSTs) were constructed and visualized using GrapeTree (https://achtman-lab.github.io/GrapeTree/MSTree_holder.html) based on the core-genome MLST (cgMLST). Tree nodes were positioned through dynamic rendering and node style was adjusted by fine-tuning the node size and kurtosis. Nodes were coloured by the pathotype of the isolates and node sizes were drawn proportionally to the number of isolates.

### 2.7 Testing and Optimization of Multiplex PCR assay

#### 2.7.1 mDPCR primers optimization

To optimize the multiplex PCR assay, each set of primers was individually tested in monoplex PCR reactions to amplify specific gene targets. Initially, combinations of two genes were tested, and the most efficient pair was selected for the initial double PCR. Subsequently, additional primer pairs were sequentially incorporated to form triple PCR reactions. This iterative process was repeated until the final multiplex PCR assay was successfully developed. The core targets of the assay included *rmpA, rmpA2, iucA*, *bla*_OXA-48-like_, and *bla*_NDM_, while *iroB, peg-344*, and *bla*_KPC_ were optionally included based on specific requirements.

#### 2.7.2 m-PCR cyclic condition optimization

The PCR reaction was prepared using the Qiagen Multiplex PCR kit, comprising HotStarTaq DNA polymerase, Multiplex PCR Buffer, and a Q-Solution. Each reaction was set up in a 20-μl volume, containing 10µL of mastermix, 2µL of Q solution, and 6µL of pooled primers (with 2µL of each forward and reverse primer for 5 genes combined to make a total of 20µL, to which 80µL of Nuclease-free water was added). Additionally, 2µL of DNA template was added to each reaction. The PCR amplification was carried out using a Veriti thermal cycler (Applied Biosystem Inc., Foster City, CA). Annealing temperatures ranging from 46°C to 56°C were tested to optimize the amplification conditions. Subsequently, 5µL of the reaction mixture was loaded onto a 2% agarose gel and electrophoresed at 130 V for 45 minutes to visualize the resulting amplicons.

#### 2.7.3 PCR Specificity & Sensitivity

The specificity of the m-PCR assay was evaluated using 150 clinical isolates of *K. pneumoniae* and twelve bacterial strains, including reference strains of *K. quasipenumoniae **ATCC 700603**, Klebsiella pneumoniae **ATCC BAA-1705*** and ***BAA-1706***, *E. coli **ATCC 35218**, P. aeruginosa **ATCC 27853**, A. baumannii **ATCC 19606**, S. aureus **ATCC 43300***, and *S. pneumoniae **ATCC 17815*** as well as strains of *Salmonella* Typhi, *Shigella* sp., *Morgenella morganii*, *Serratia marcescens,* and *Proteus mirabilis* from IHMA. Genomic DNA extraction was carried out following the procedure outlined in Section 2.5, and PCR amplification was performed as described in Section 2.7.2.

A tenfold serial dilution procedure was adapted to assess the sensitivity of the m-PCR assay. Initially, the positive control strain (B1644-Accession ID-GCA_047922575) was cultured on MacConkey agar, and a single colony was inoculated into Muller Hilton Broth, followed by adjustment to 0.5 McFarland standard. The suspension was serially diluted in sterile saline from 10^-1^ to 10^-8^, and 10 µL from each dilution was plated on MacConkey agar to determine the CFU/mL after 24-hour incubation at 37°C. Additionally, 200 µL from each dilution was used for DNA extraction, followed by m-PCR amplification to detect target genes. The experiment, performed in duplicate, determined sensitivity by consistently identifying the lowest dilution (CFU/mL).

These DNA samples served as templates for PCR amplification. Further, the DNA was serially diluted to achieve concentrations ranging approximately from 50 ng/L to 70 pg/L. Subsequently, 2µL of each dilution was employed as a template to evaluate the sensitive

## Results

### 3.1 Screening of gene targets

A total of 12,846 genomes were identified as hvKp through a comprehensive screening of the global collection in the NCBI Pathogen Detection Database (*Table S2*). The identification criteria included the presence of key virulence genes such as *rmpA*, *rmpA2*, and *iucA*, either individually or in combination. Analysis using the Kleborate tool demonstrated the distribution of key virulence markers among the identified *K. pneumoniae* genomes. The *iucA* gene was the most prevalent, detected in 96.32% (12,374/12,846) of genomes, followed by *peg-344* in 73.9% (9495/12,846), *rmpA2* in 70% (8992/12,846), *rmpA* in 48.25% (6199/12,846) and *iroB* in 30.83% (3961/12,846). Additionally, 68.72% (8828/12,846) of the genomes carried carbapenemase genes. Among these, *bla*_KPC_ variants were the most common (4,253/8828), followed by *bla*_NDM_ (2,652/8828) and *bla*_OXA-48-like_ (2,429/8828).

A subset analysis of genomes from Indian clinical settings revealed a similar trend in virulence determinants but notable differences in resistance profiles. In this dataset, *iucA* was detected in 99.6% (523/525) of genomes, while *rmpA was in 16% (87/525), rmpA2* was present in 36% (189/525), *iroB* in 16% (84/525), and *peg-344* in 38.09% (200/525). However, the resistance landscape differed: 76.95% (404/525) of Indian genomes carried carbapenemase genes, predominantly *bla*_OXA-48-like_ (93.81%, 379/404), followed by *bla*_NDM_ (10.89%, n=44/404), with some percentage among them having both the genes and four isolates carrying *bla*_NDM_ co-carried *bla*_KPC_ gene were detected.

### 3.2 Primer design and *in silico* validation

We selected five candidate genes (*rmpA, rmpA2, iucA and bla*_NDM_, *bla*_OXA-48-like_) to differentiate CR-hvKp based on their prevalence in Indian clinical settings. Primers for *iroB* and *peg-344* were designed but not included in the primary panel, as these genes can be used in the panel on specific requirements. Similarly, *bla*_KPC_ is rarely found in India isolates, primers were included only for standardization purposes. All primers were designed to target conserved regions of hypervirulence-associated genes (*rmpA, rmpA2, iucA, iroB, peg-344*) and resistance genes (*bla*_NDM_, *bla*_OXA-48-like_ and *bla*_KPC_ variants).

Designed primers were optimized to have a GC content near 50%, a ΔTm < 2°C, and a length of 18–22 bases. Evaluations using *in silico* PCR confirmed high specificity for target sequences and no significant off-target amplification. The designed primers generated *in silico* amplification products of *rmpA (530 bp)*, *rmpA2 (470 bp)* and *iucA (170 bp), iroB (219 bp), peg-344 (313 bp)* in hypervirulent reference strains such as *K. pneumoniae* KCTC 2242 (NC_017540) and *K. pneumoniae* NTUH-K2044 (NC_012731). The primers for *bla*_NDM_, *bla*_OXA-48-like,_ and *bla*_KPC_ produced amplicons of *401 bp, 630 bp*, and *798 bp* respectively. The detailed list of all primers is mentioned in **Table 1**.

### 3.3 Bacterial Strain and Phenotypic Characterization

In this study, 150 clinical isolates of *K. pneumoniae* associated with bacteremia or respiratory infections were randomly selected for primer evaluation and phylogenomic analysis. Following confirmation of these isolates as *K. pneumoniae*, they were assessed for hmv using the string test, which identified 22% (n=33/150) as hmv-positive. Additionally, the isolates exhibited significant antimicrobial resistance, with only 30% (n=45/150) showing susceptibility to carbapenems, specifically ertapenem and meropenem.

### 3.4 Genome-based Population Structure of Bacterial Strains

The minimum spanning tree (MST) of the *K. pneumoniae* study isolates reveals a diverse population structure, with clear clustering based on sequence types (STs). The predominant clone was ST2096, which comprised 26.7%, (40/150) isolates, indicating its dominance within the studied population. ST147, the second largest cluster with 21.3% (32/150) isolates, is a globally recognized high-risk clone associated with carbapenem resistance. Another prominent cluster, ST231, associated with 13.3% of isolates, is notable for its strong association with antimicrobial resistance genes in Indian clinical settings. Hypervirulent clones, including ST23 and ST65, were observed in smaller clusters with 4% (6/150) and 1.3% (2/150) isolates, respectively. Additionally, the population demonstrates considerable genetic diversity, with smaller clusters of sequence types (STs) such as ST101, ST14, ST45, and ST16. The phylogenetic relationships inferred from the MST highlight the coexistence of multidrug-resistant (MDR) and hypervirulent clones, with evidence of convergence in certain lineages. Based on Kleborate’s classification, 58% (87/150) of the isolates were identified as MDR-hvKp. In contrast, 19% (28/150) of the isolates were hypervirulent but lacked extended-spectrum beta-lactamases (ESBLs) or carbapenemase genes, while the remaining 23% (35/150) carried low-virulence features such as yersiniabactin, with no ESBL or carbapenemase genes, and were classified as classical *K. pneumoniae* (cKp).

The minimum spanning tree (MST) based on cgMLST revealed the clonal distribution of *Klebsiella pneumoniae* isolates. The predominant sequence types were ST2096 (40 isolates), ST147 (32 isolates), and ST231 (20 isolates), forming major clusters that were centrally positioned, linking multiple other STs, suggesting their epidemiological significance. Other notable STs included ST23, ST15, ST16, ST395, and ST420 (5–6 isolates each) **Figure 1**.

**Figure 1:**
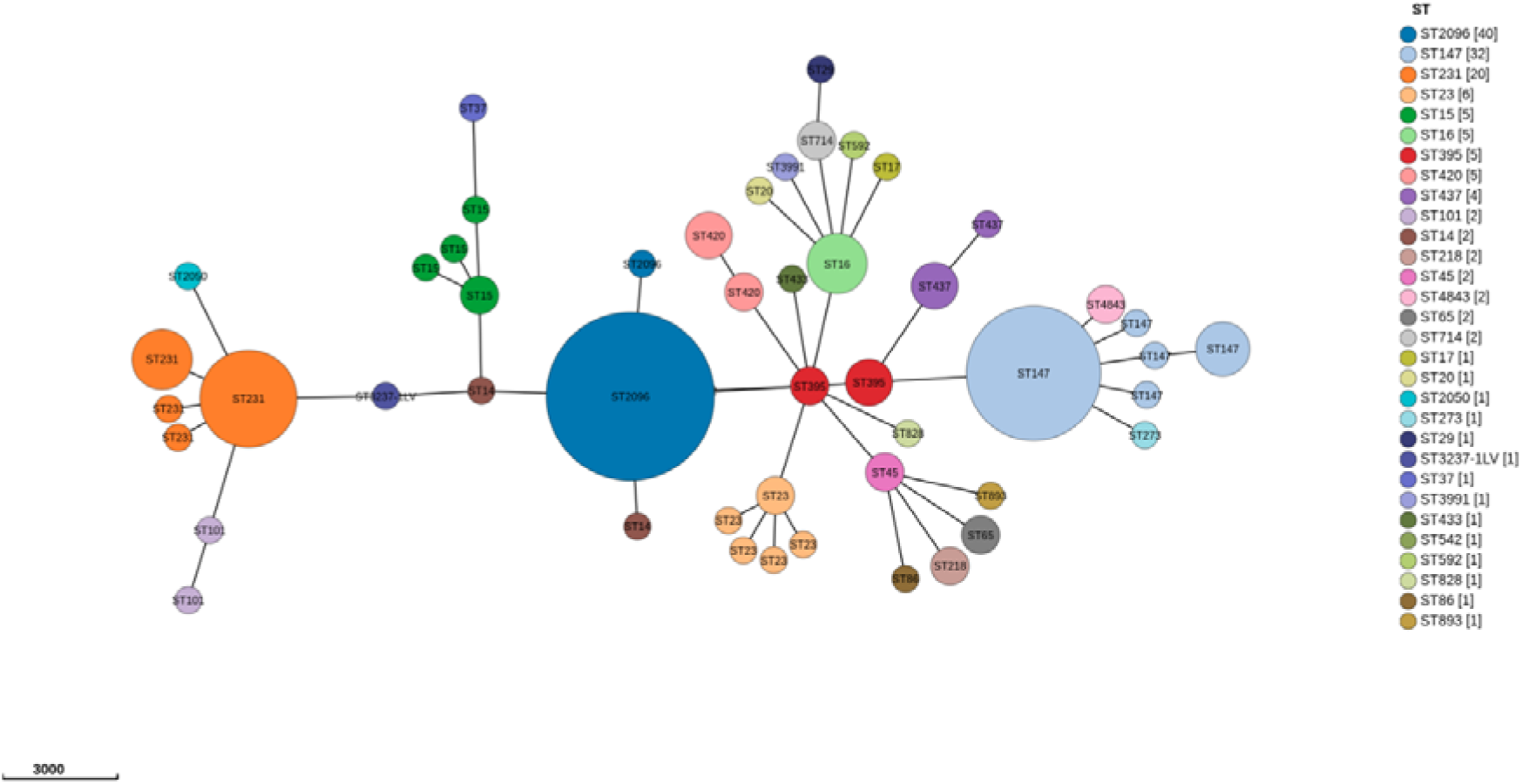
Minimum spanning tree (MST) for n=150 isolates based on cgMLST. The tree was generated using GrapeTree, with nodes representing different sequence types (STs). Node sizes are proportional to the number of isolates within each ST, and colors differentiate sequence types. The scale represents the genetic distance.

### 3.5 Optimization of PCR Conditions

The multiplex PCR assay was optimized to detect five target genes (*iucA, bla_NDM_, rmpA, rmpA2,* and bla*_OXA-48-like_*) through a stepwise approach, progressing from a duplex to a pentaplex assay. The optimal annealing temperature was determined to be 50°C, with 35 cycles ensuring successful amplification. This condition provided consistent and reliable results for all target genes when multiplexed (***Supplementary Figure c***). For all the optimization experiments positive control B1644-Accession ID-GCA_047922575 was used.

Additionally, markers such as *iroB*, *peg-344* and *bla*_KPC_ were optimized as an extended version of the multiplex assay. The detailed cyclic conditions for all the markers are mentioned in ***Table 2***. All eight target genes were also amplified in singleplex PCR, confirming the functionality of individual primers. The multiplex PCR assay also demonstrated successful amplification of the selected markers shown in ***Supplementary Figure d.*** Agarose gel electrophoresis confirmed the successful amplification of target genes in the multiplex PCR assay. Various combinations of genes were tested to ensure optimal amplification without cross-reactivity (***Supplementary Figure e)***. Distinct bands correspond to *iucA, rmpA, rmpA2, bla*_NDM_, and *bla*_OXA-48-like_, validating the specificity of the assay as observed in **Figure 2**.

**Figure 2:**
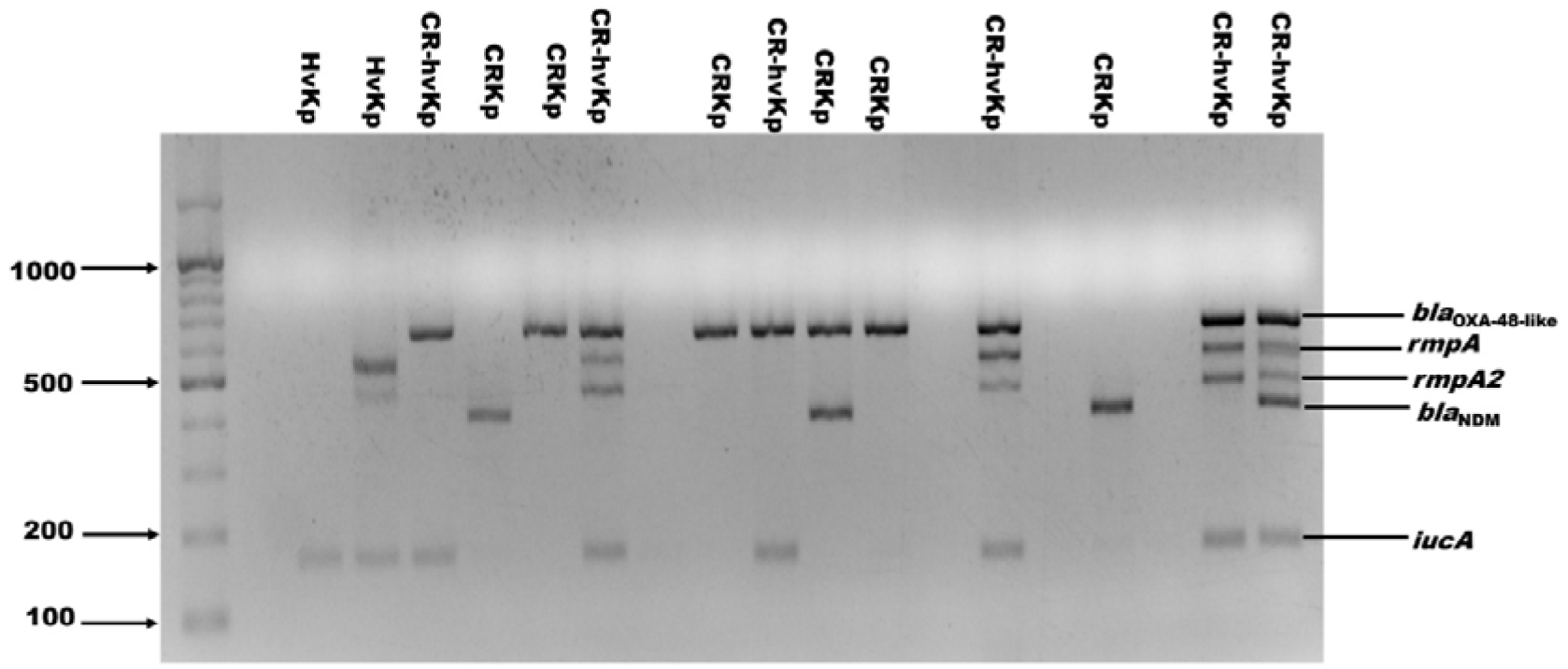
Multiplex PCR detection of resistance and virulence genes in Klebsiella pneumoniae pathotypes. Lanes show amplification of rmpA, rmpA2, iucA, bla_NDM_, and bla_OXA-48-like_ genes in various K. pneumoniae isolates. A 100 bp DNA ladder is shown in the first lane. The presence of multiple bands indicates convergence of resistance and virulence traits.

**Table 2:** Cycling conditions (Qiagen Multiplex) Table 1: Cycling conditions for the detection of virulence (rmpA, rmpA2, iucA, iroB, peg-344) and resistance (blaNDM, blaOXA-48-like, blaKPC) genes, including annealing temperatures, cycle numbers, and extension times for both the primary multiplex assay and the extended panel.

|  | Temperature (°C) | Time | Cycles |
| --- | --- | --- | --- |
| **Initial Denaturation** | 95 | 10 mins | 35 |
| **Denaturation** | 94 | 30 sec |  |
| **Annealing** | 50 (Works for all 8 genes) | 1 |  |
| **Extension** | 72 | 1 min |  |
| **Final Extension** | 72 | 10 mins |  |
| **Hold** | 4 | ∞ |  |

### 3.6 Sensitivity and Specificity

The assay demonstrated 100% specificity, with no amplification observed in non-target bacterial strains. Sensitivity tests revealed a consistent detection limit of 1 ng/µL for the target genes across triplicate experiments (***Supplementary figure a, b)***. Amplification results confirmed the presence of virulence genes (*rmpA, rmpA2, iucA*) and resistance genes (*bla*_NDM_, *bla*_OXA-48-like_), with successful detection up to a bacterial load corresponding to a 10[² dilution (525 CFU/mL). For standardization positive control B1644-Accession ID-GCA_047922575 was used.

To assess the feasibility of detecting multiple targets in a single reaction, we tested different gene combinations in multiplex PCR assays. Various sets of hypervirulence (*rmpA, rmpA2, iucA, iroB, peg-344*) and resistance markers (*bla*_NDM_*, bla_OXA-48-like_, bla_KPC_*) were evaluated to determine compatibility and amplification efficiency. The optimized pentaplex assay (*iucA, bla_NDM_, rmpA, rmpA2*, and *bla*_OXA-48-like_) consistently yielded distinct and reproducible bands. Additional genes, including *iroB, peg-344,* and *bla*_KPC_, were incorporated in extended multiplex reactions, demonstrating successful amplification without cross-reactivity. These results confirm the adaptability of the assay for targeted applications, allowing for tailored detection based on specific diagnostic needs as shown in ***Supplementary Figure f***.

### 3.7 Phenotypic and PCR Correlation of Hypervirulence and Carbapenem Resistance

The correlation between the hmv phenotype and virulence markers is key to assessing the string test’s diagnostic value. Among 150 isolates, the string test identified hmv in 33, with 79% (26/33) carrying at least one of three PCR-detected virulence markers. However, 76% (88/116) of hmv-negative isolates also harbored at least one hypervirulence-associated marker (Table S1). These results suggest that relying solely on the string test risks false negatives, underscoring the need for molecular diagnostics to identify hvKp accurately. In contrast, resistance phenotypes showed perfect correlation with genotypic findings: all 105 phenotypically carbapenem-resistant isolates carried either *bla*_OXA-48-like_ (59/105), *bla*_NDM_ (21/105), or both genes (25/105), while none of the carbapenem-susceptible strains tested positive for these genes by PCR.

### 3.8 WGS and PCR Correlation of Hypervirulence and Carbapenem Resistance

WGS and multiplex PCR analysis demonstrated a 100% correlation in detecting hypervirulence and carbapenem resistance determinants. All isolates carrying key virulence genes (*iucA, rmpA, rmpA2*) in WGS were also confirmed by PCR, reinforcing the reliability of the selected molecular markers. Similarly, carbapenem-resistant isolates harbored *bla*_OXA-48-like_, *bla*_NDM_, or both, with perfect concordance between WGS and PCR results. No discrepancies were observed, indicating that the developed multiplex PCR-based screening effectively detects hypervirulence and resistance markers with the same accuracy as WGS.

## 4 Discussion

Healthcare-associated infections (HAIs) caused by *K. pneumoniae* (Kp) remain a critical global challenge due to their high prevalence, widespread antimicrobial resistance, and diverse virulence characteristics (1,9). The increasing incidence of CRKp strains has further complicated this issue, as many of these strains carry virulence-associated genes (*rmpA, rmpA2, iucA, iroB and peg-344*) alongside resistance determinants, resulting in severe and often untreatable infections (15, 32). Complicating their detection, many of these convergent clones, which exhibit both multidrug resistance and hypervirulence traits, lack the hypermucoviscous phenotype traditionally assessed using the string test. This limitation significantly undermines the utility of phenotypic methods in accurately identifying these high-risk strains. To address these diagnostic challenges, we have attempted to systematically screen global hvKp genomes and developed a robust m-PCR assay to accurately identify CR-hvKp, addressing key gaps in existing diagnostic approaches.

Genome analysis of ∼12000 hvKp genomes from the NCBI Pathogen Detection Database revealed a high prevalence of hypervirulence-associated genes, with *iucA* (aerobactin) being the most frequently detected, followed by *peg-344/rmpA2*. While less common, other hypervirulence markers such as *iroB* and *rmpA* may also be useful depending on their regional distribution. Carbapenemase genes were identified in 69.17% of the genomes, with *bla*_KPC_ being the most prevalent globally. However, analysis of Indian clinical isolates demonstrated a predominance of *bla*_OXA-48-like_ genes, followed by *bla*_NDM_, highlighting significant regional differences in resistance mechanisms. These findings are consistent with previous reports indicating that *bla*_KPC_ is the dominant carbapenemase globally, whereas carbapenem resistance in *K. pneumoniae* in India is often associated with endemic clones harboring *bla*_OXA-48-like_ and *bla*_NDM_ (33–35). Together, these genes serve as critical markers for both virulence and drug resistance, enabling comprehensive pathotype classification of clinical isolates. Minimum spanning tree analysis of our 150 clinical isolates identified distinct clones circulating in India, including ST2096, ST147, and ST231, which differ from the globally dominant hospital-associated clones. Notably, 58% of these isolates were classified as MDR-hvKp, indicating a high convergence rate that exceeds Southeast Asian estimates (20–30%) (16). This high prevalence of MDR-hvKp in India underscores the clinical relevance of this mPCR assay in this setting. Given the regional differences in clone distribution and resistome profiles, we have selected five key genetic markers that serve as critical indicators of both virulence and drug resistance, enabling comprehensive pathotype classification of clinical isolates.

The significance of this assay lies in its ability to simultaneously detect key virulence and resistance determinants within a single reaction, thereby enhancing diagnostic efficiency. By eliminating the need for sequential testing or the resource-intensive process of WGS, this approach significantly reduces turnaround time while ensuring rapid and accurate pathogen characterization. Among the 150 clinical isolates tested, the assay demonstrated 100% concordance with WGS for *iucA, rmpA, rmpA2, bla*_NDM_, and *bla*_OXA-48-like_, with a detection sensitivity of 1 ng/µL DNA or 525 CFU/mL. This performance far exceeds that of the string test, which identified hmv in only 33 of 150 isolates, failing to detect another 88 isolates that harboured virulence markers. These findings underscore the limitations of phenotypic screening for hypervirulence. Such limitations, consistent with the findings of Russo et al. (2018) (23) and Zhu et al. (2021) (24), underscore the unreliability of phenotypic tests for hvKp detection, particularly in cases where mutations in *rmpA/rmpA2* or alternative virulence factors like *peg-344* drive pathogenicity (27, 36). In contrast, all 105 carbapenem-resistant isolates exhibited perfect phenotypic-genotypic correlation, reaffirming the assay’s utility for CRKp detection. The ability to rapidly and accurately identify CR-hvKp could significantly reduce mortality rates, which often exceed 50% in delayed-treatment cases (15), and enhance antimicrobial stewardship, particularly in high-prevalence settings such as India, where 57% of *K. pneumoniae* bloodstream infections are carbapenem-resistant (10).

Previous studies, such as those by Fangyou et al. (2018), have attempted to differentiate carbapenem-resistant from hypervirulent *K. pneumoniae* strains using multiplex PCR assays (37). While they developed three sets of multiplex primers wherein the first set of primers was designed specifically for certain sequence types (ST11/258, ST23, ST86, ST65 and ST375), the second set was for capsular polymerase genes specific to various K types like K1, K2, KL64 and KL47(37). Similarly, another study developed a multiplex PCR assay targeting seven virulence genes (*rmpA, allS, kfu, iuc, iro, fimH, uge*) along with K1/K2 capsular serotypes, enhancing the detection of hypervirulent strains (38). More recently, LAMP and multiplex qRT-PCR assays offer improved sensitivity and specificity for detecting hypervirulent *K. pneumoniae* (39). While these assays contribute to rapid and specific identification, the m-PCR approach is confined to predefined virulence genes and capsular types, potentially overlooking emerging genetic variants and converging traits. A previous study from our setting (26) prioritized *iucA* as the sole marker for hvKp detection. However, our inclusion of multiple virulence markers addresses variability in *rmpA/rmpA2*, as highlighted by Lin et al. (2020) (27). Our assay, in contrast, provides a more comprehensive solution by simultaneously detecting both virulence and resistance factors, overcoming the limitation of focusing on a narrow set of markers. Our assay is designed to work effectively within the Indian subcontinent’s genomic diversity, including prevalent sequence types like ST2096, ST147, and ST231, which are not commonly identified in studies from regions such as the United States. This region-specific focus is essential for the accurate diagnosis of *K. pneumoniae* infections and underscores the importance of tailored diagnostic tools.

A key strength of our m-PCR assay is its comprehensive detection capability, enabling the simultaneous identification of all three major *K. pneumoniae* pathotypes (CRKp, hvKp, and CR-hvKp). Unlike the string test, which is subjective and prone to false negatives, this molecular approach accurately detects both virulence and resistance determinants, ensuring earlier diagnosis and intervention—critical for preventing severe hvKp infections. Additionally, the assay’s high specificity ensures that only *K. pneumoniae* genes are amplified, eliminating the risk of cross-reactivity with other bacterial species. To further enhance its clinical utility, we have incorporated regionally relevant targets, optimizing five core genes (*iucA, rmpA, rmpA2*, *bla*_NDM_, *bla*_OXA-48-like_) while also allowing for the inclusion of *peg-344*, *iroB*, and *bla*_KPC_ based on epidemiological needs. These adaptations improve the global applicability of the assay. Compared to WGS and single-plex PCR, which are expensive and time-consuming, this m-PCR assay offers a cost-effective, high-throughput solution suitable for routine diagnostics, particularly in resource-limited settings. Since this assay has been validated on diverse clinical isolates across India, it offers a high-throughput, reliable alternative to traditional diagnostic methods, making it especially valuable for clinical diagnostics in Indian hospitals and similar global settings.

Despite these strengths, there are limitations to the multiplex PCR assay. Although it covers a broad spectrum of prevalent genes to differentiate between the three pathotypes of *K. pneumoniae* strains, it does not target other virulence factors like those involved in biofilm formation.

In conclusion, this study presents the development and validation of a multiplex PCR assay that offers a significant improvement over existing methods for detecting *Klebsiella pneumoniae* pathotypes in clinical isolates. By simultaneously detecting virulence and resistance genes, the assay provides a comprehensive diagnostic tool that is suitable for rapid clinical implementation. Beyond clinical diagnostics, this assay enhances molecular surveillance and outbreak investigations by enabling real-time tracking of strain prevalence. Its ability to rapidly and accurately identify carbapenem-resistant and hypervirulent *K. pneumoniae* supports infection control efforts, targeted interventions, and antimicrobial stewardship in both healthcare and community settings. Overall, this assay provides a valuable solution for managing *K. pneumoniae* infections in India and similar high-burden regions, addressing the growing challenge of antimicrobial resistance and hypervirulence through efficient and accessible molecular diagnostics.

## 7. Ethical Approval

The study was approved by the Institutional Review Board (IRB) Christian Medical College, Vellore, min. no. 15248 dated 22.03.2023.

## 9. Authors contribution

**Sanika Mahesh Kulkarni**□ – Conceptualization, Data Curation, Methodology, Investigation, Formal analysis, Writing – original draft, and Visualization. **Jobin John Jacob**□-Data curation, Validation and Writing – review & editing. **Praveen T**□-Methodology and Formal analysis. **Aravind V**□ –Methodology and Software. **Subbulakshmi R**□-Data curation and Software. **Preethi S**□-Methodology and Formal analysis. **Binesh Lala Y**-Supervision and Writing – review & editing. **Karthik Gunasekaran**□ – Investigation and Formal Analysis. **Abi Manesh**□

Supervision and Clinical oversight. **Shraddha M Karve**□ – Writing – review & editing. **Sudarsana J**□, **Sanjay Bhattacharya**□, **Anand Shah**□, **Savitha Nagaraj**□, **Priyadarshini Padaki**□, **Jayakumar S**□, **Renu Mathew**□, **Rudresh SM**□, **Shariqa Qureshi**□, **S Nivedhana**□, and **Geethu Joe**^n^ – Resources and Validation. **Ekadashi Rajni**□-Writing – review & editing. **Kamini Walia**□ – Project administration, Funding acquisition, Writing – review & editing and Supervision. **Balaji Veeraraghavan**□* –Conceptualization, Supervision, Project administration, Funding acquisition, and Writing – review & editing

## 9. Acknowledgements

The authors thank the Department of Clinical Microbiology, Christian Medical College and Hospital, Vellore, for providing us with all the necessary facilities to conduct our study. I would also like to mention that this work is a part of my Ph.D. Thesis of **THE TAMIL NADU Dr. M.G.R. MEDICAL UNIVERSITY.**

## Supporting information

Supplementary Figure a-f, Agarose gel electrophoresis images showing validation and sensitivity of the m-PCR

## Data Availability

All data produced in the present study are available upon reasonable request to the authors

## Supplementary Figures

**Supplementary Figure.**
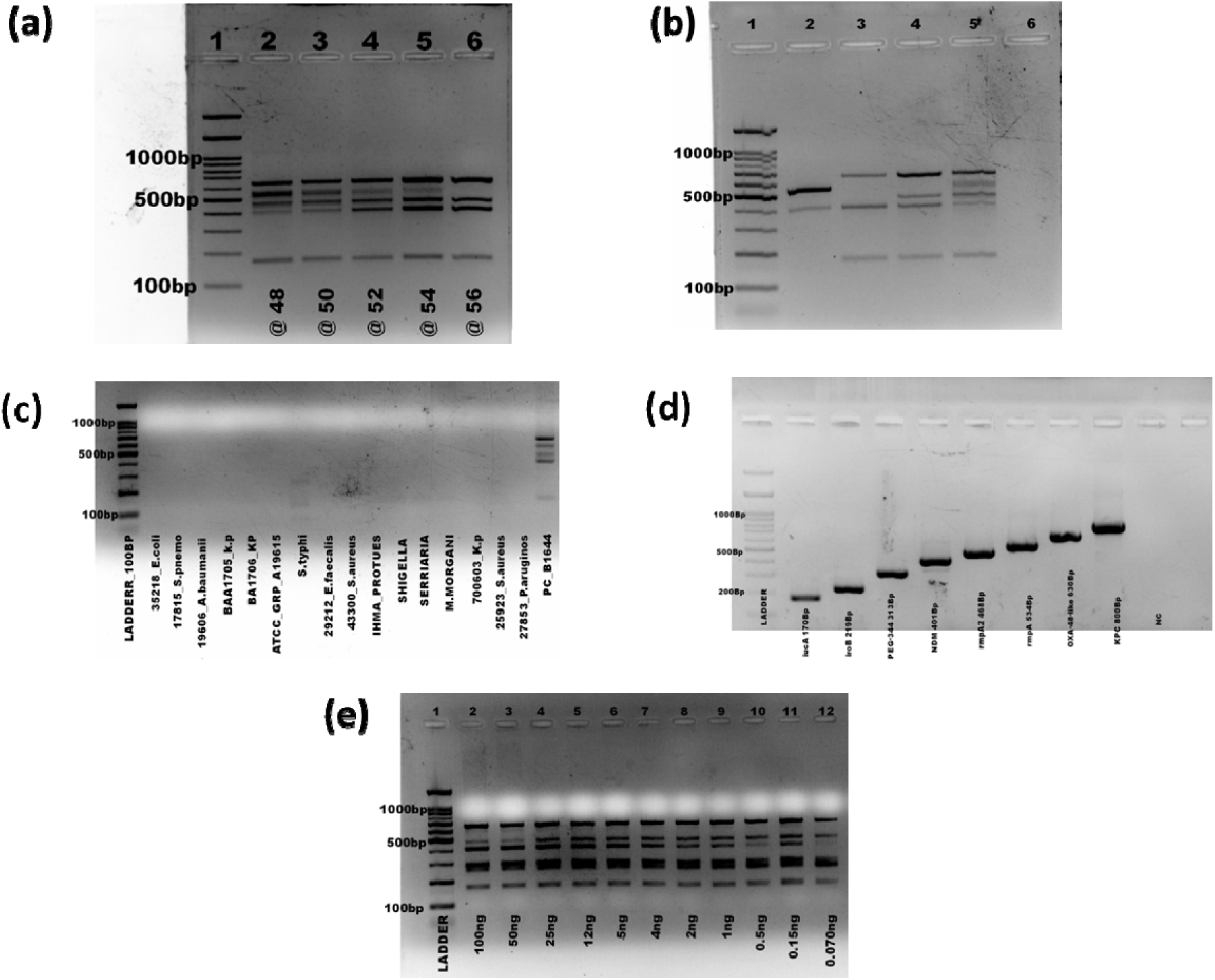
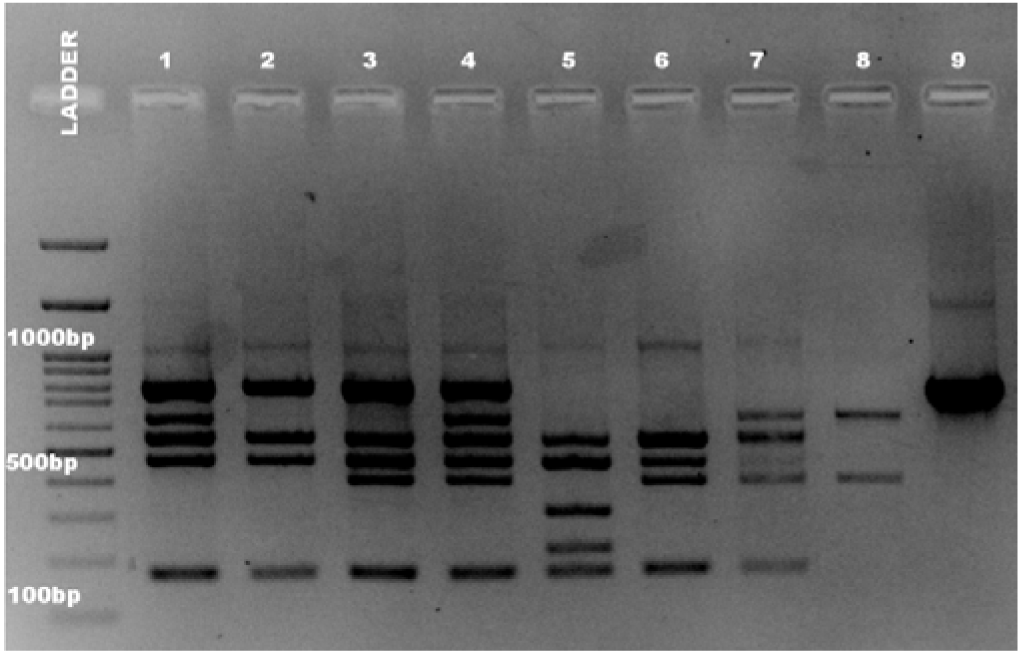
**a**. Determination of annealing temperature of the multiplex PCR. Lane 1: 1000 bp DNA marker; Lane 1: 48°C annealing; Lane 2: 50°C annealing; Lane 3: 52°C annealing; Lane 4: 54°C annealing; Lane 5:56°C annealing. **Figure b.** Optimization of the primers of m-PCR. Lane 1: 1000 bp DNA marker; Lane 1: duplex PCR; Lane 2: triplex PCR; Lane 3: quadruplex PCR; Lane 4: pentaplex PCR formed by *iucA, bla_NDM_*, *rmpA, rmpA2* and *bla_OXA-4-like_*. **Figure c**. Determination of specificity of the multiplex PCR. Lane 1: 1000 bp DNA marker, Lane 2-16: the template 14 bacterial strains of m-PCR which are negative. Lane 17: positive control. **Figure d.** Amplification of target gene of the multiplex PCR. Lane 1: 1000 bp DNA marker; Lanes 2–9: the template of single genes amplification respectively for *iucA, iroB, peg-344, bla_NDM_, rmpA2, rmpA*, *bla_OXA-48-like_ and bla_KPC._* **Figure e**. Determination of the sensitivity of the multiplex PCR for bacterial genomic DNA detection for strain B1644, Lane 1: 1000 bp DNA marker; a Lanes 2–12: the concentration of Klebsiella pneumoniae DNA were 100 ng, 50 ng, 25 ng, 12 ng, 5 ng, 4 ng, 1 ng, 500 pg, 150 pg, and 70 pg respectively. *Figure f.* We evaluated various multiplex PCR combinations to determine the optimal setup for detecting key virulence (*rmpA, rmpA2, iucA, iroB, peg-344*) and resistance (*bla_NDM_, bla_OXA-48-like_, blaKPC*) genes. Different gene combinations were tested to assess their compatibility within a single assay. Lane-wise, the results are as follows: the first lane contains the molecular size marker (ladder), followed by multiplex amplification of *rmpA, rmpA2, iucA, bla_OXA-48-like_, bla_KPC_* (Lane 1), *rmpA, rmpA2, iucA, bla_KPC_, bla_NDM_*(Lane 2), and *rmpA, rmpA2, iucA, bla_KPC_, bla_NDM_, bla_OXA-48-like_* (Lane 3). Lane 4 includes the extended panel with *rmpA, rmpA2, iucA, iroB, and peg-344*, while Lane 5 and Lane 6 represent multiplex combinations with *rmpA, rmpA2, iucA, bla_NDM_* and *rmpA, rmpA2, iucA, bla_NDM_, bla_OXA-48-like_*, respectively. Lane 7 shows amplification of *bla_OXA-48-like_*and *bla_NDM_* alone, whereas Lane 8 confirms the presence of *blaKPC* individually. These results highlight the feasibility of a robust multiplex PCR approach for detecting hypervirulence and carbapenem resistance markers in *Klebsiella pneumoniae*, with additional genes available for extended analysis as needed.

## References

1. Shon AS, Bajwa RPS, Russo TA. Hypervirulent (hypermucoviscous) Klebsiella pneumoniae. Virulence. 2013 Feb 15;4(2):107–18.

2. Catalán-Nájera JC, Garza-Ramos U, Barrios-Camacho H. Hypervirulence and hypermucoviscosity: Two different but complementary Klebsiella spp. phenotypes? Virulence. 2017 Oct 3;8(7):1111–23.

3. Gorrie CL, Mirceta M, Wick RR, Edwards DJ, Thomson NR, Strugnell RA, et al. Gastrointestinal Carriage Is a Major Reservoir of Klebsiella pneumoniae Infection in Intensive Care Patients. Clin Infect Dis. 2017 Jul 15;65(2):208–15.

4. Martin RM, Cao J, Brisse S, Passet V, Wu W, Zhao L, et al. Molecular Epidemiology of Colonizing and Infecting Isolates of Klebsiella pneumoniae. mSphere. 2016 Oct 19;1(5):e00261–16.

5. Meatherall BL, Gregson D, Ross T, Pitout JDD, Laupland KB. Incidence, risk factors, and outcomes of Klebsiella pneumoniae bacteremia. Am J Med. 2009 Sep;122(9):866–73.

6. Podschun R, Ullmann U. Klebsiella spp. as nosocomial pathogens: epidemiology, taxonomy, typing methods, and pathogenicity factors. Clin Microbiol Rev. 1998 Oct;11(4):589–603.

7. Russo TA, Marr CM. Hypervirulent Klebsiella pneumoniae. Clin Microbiol Rev. 2019 Jun 19;32(3):e00001–19.

8. Dong N, Yang X, Chan EWC, Zhang R, Chen S. Klebsiella species: Taxonomy, hypervirulence and multidrug resistance. eBioMedicine [Internet]. 2022 May 1;79. Available from: https://www.thelancet.com/journals/ebiom/article/PIIS2352-3964(22)00182-7/fulltext#seccesectitle0005

9. Navon-Venezia S, Kondratyeva K, Carattoli A. Klebsiella pneumoniae: a major worldwide source and shuttle for antibiotic resistance. FEMS Microbiology Reviews. 2017 May 1;41(3):252–75.

10. AMR_Annual_Report_2021.pdf [Internet]. Available from: https://main.icmr.nic.in/sites/default/files/upload_documents/AMR_Annual_Report_2021.pdf

11. WHO publishes list of bacteria for which new antibiotics are urgently needed [Internet]. [cited 2023 Jan 27]. Available from: https://www.who.int/news/item/27-02-2017-who-publishes-list-of-bacteria-for-which-new-antibiotics-are-urgently-needed

12. Paczosa MK, Mecsas J. Klebsiella pneumoniae: Going on the Offense with a Strong Defense. Microbiol Mol Biol Rev. 2016 Jun 15;80(3):629–61.

13. Wang X, Zhao J, Ji F, Chang H, Qin J, Zhang C, et al. Multiple-Replicon Resistance Plasmids of Klebsiella Mediate Extensive Dissemination of Antimicrobial Genes. Front Microbiol. 2021 Oct 27;12:754931.

14. Zhang R, Lin D, Chan EW chi, Gu D, Chen GX, Chen S. Emergence of Carbapenem-Resistant Serotype K1 Hypervirulent Klebsiella pneumoniae Strains in China. Antimicrob Agents Chemother. 2015 Dec 31;60(1):709–11.

15. Gu D, Dong N, Zheng Z, Lin D, Huang M, Wang L, et al. A fatal outbreak of ST11 carbapenem-resistant hypervirulent Klebsiella pneumoniae in a Chinese hospital: a molecular epidemiological study. Lancet Infect Dis. 2018 Jan;18(1):37–46.

16. Wyres KL, Nguyen TNT, Lam MMC, Judd LM, van Vinh Chau N, Dance DAB, et al. Genomic surveillance for hypervirulence and multi-drug resistance in invasive Klebsiella pneumoniae from South and Southeast Asia. Genome Medicine. 2020 Jan 16;12(1):11.

17. Chen L, Kreiswirth BN. Convergence of carbapenem-resistance and hypervirulence in Klebsiella pneumoniae. Lancet Infect Dis. 2018 Jan;18(1):2–3.

18. Raj S, Sharma T, Pradhan D, Tyagi S, Gautam H, Singh H, et al. Comparative Analysis of Clinical and Genomic Characteristics of Hypervirulent Klebsiella pneumoniae from Hospital and Community Settings: Experience from a Tertiary Healthcare Center in India. Microbiol Spectr. 2022 Oct 26;10(5):e0037622.

19. Vandhana V, Saralaya KV, Bhat S, Shenoy Mulki S, Bhat AK. Characterization of Hypervirulent Klebsiella pneumoniae (Hv-Kp): Correlation of Virulence with Antimicrobial Susceptibility. Int J Microbiol. 2022;2022:4532707.

20. Banerjee T, Wangkheimayum J, Sharma S, Kumar A, Bhattacharjee A. Extensively Drug-Resistant Hypervirulent Klebsiella pneumoniae From a Series of Neonatal Sepsis in a Tertiary Care Hospital, India. Front Med (Lausanne). 2021;8:645955.

21. Mukherjee S, Mitra S, Dutta S, Basu S. Neonatal Sepsis: The Impact of Carbapenem-Resistant and Hypervirulent Klebsiella pneumoniae. Front Med (Lausanne). 2021;8:634349.

22. Yadav B, Mohanty S, Behera B. Occurrence and Genomic Characteristics of Hypervirulent Klebsiella pneumoniae in a Tertiary Care Hospital, Eastern India. Infect Drug Resist. 2023;16:2191–201.

23. Russo TA, Olson R, Fang CT, Stoesser N, Miller M, MacDonald U, et al. Identification of Biomarkers for Differentiation of Hypervirulent Klebsiella pneumoniae from Classical K. pneumoniae. J Clin Microbiol. 2018 Aug 27;56(9):e00776–18.

24. Zhu J, Wang T, Chen L, Du H. Virulence Factors in Hypervirulent Klebsiella pneumoniae. Front Microbiol. 2021 Apr 8;12:642484.

25. Yan C, Zhou Y, Du S, Du B, Zhao H, Feng Y, et al. Recombinase-Aided Amplification Assay for Rapid Detection of Hypervirulent Klebsiella pneumoniae (hvKp) and Characterization of the hvKp Pathotype. Microbiology Spectrum. 2023 Mar 13;11(2):e03984–22.

26. Shankar C, Basu S, Lal B, Shanmugam S, Vasudevan K, Mathur P, et al. Aerobactin Seems To Be a Promising Marker Compared With Unstable RmpA2 for the Identification of Hypervirulent Carbapenem-Resistant Klebsiella pneumoniae: In Silico and In Vitro Evidence. Frontiers in Cellular and Infection Microbiology [Internet]. 2021 [cited 2022 Dec 16]; 11. Available from: https://www.frontiersin.org/articles/10.3389/fcimb.2021.709681

27. Lin Z wei, Zheng J xin, Bai B, Xu G jian, Lin F jun, Chen Z, et al. Characteristics of Hypervirulent Klebsiella pneumoniae: Does Low Expression of rmpA Contribute to the Absence of Hypervirulence? Front Microbiol. 2020 Mar 17;11:436.

28. Yu WL, Lee MF, Chang MC, Chuang YC. Intrapersonal mutation of rmpA and rmpA2: A reason for negative hypermucoviscosity phenotype and low virulence of rmpA-positive Klebsiella pneumoniae isolates. Journal of Global Antimicrobial Resistance. 2015 Jun 1;3(2):137–41.

29. Poirel L, Walsh TR, Cuvillier V, Nordmann P. Multiplex PCR for detection of acquired carbapenemase genes. Diagn Microbiol Infect Dis. 2011;70(1):119–123. doi:10.1016/j.diagmicrobio.2010.12.002

30. Clinical and Laboratory Standards Institute (CLSI). Performance standards for antimicrobial susceptibility testing. CLSI supplement M100, 32nd ed. Wayne, PA: CLSI; 2022.

31. Clinical and Laboratory Standards Institute (CLSI). Performance standards for antimicrobial susceptibility testing. CLSI supplement M100, 33rd ed. Wayne, PA: CLSI; 2023.

32. Zhan L, Wang S, Guo Y, Jin Y, Duan J, Hao Z, et al. Outbreak by hypermucoviscous *Klebsiella pneumoniae* ST11 isolates with carbapenem resistance in a tertiary hospital in China. Front Microbiol. 2021;12:1234.

33. Mathur P, Malpiedi P, Walia K, Srikantiah P, Gupta S, Lohiya A, et al. Health-care-associated bloodstream and urinary tract infections in a network of hospitals in India: a multicentre, hospital-based, prospective surveillance study. The Lancet Global Health. 2022 Sep 1;10(9):e1317–25

34. Manesh A, Shankar C, George MM, et al. Clinical and genomic evolution of carbapenem-resistant Klebsiella pneumoniae bloodstream infections over two time periods at a tertiary care hospital in South India: a prospective cohort study. Infect Dis Ther. 2023;12:1319–1335. doi:10.1007/s40121-023-00803-3.

35. Shankar C, Kumar S, Venkatesan M, Veeraraghavan B. Emergence of ST147 Klebsiella pneumoniae carrying blaNDM-7 on IncA/C2 with ompK35 and ompK36 mutations in India. J Infect Public Health. 2019;12(5):741–743.

36 Bulger J, MacDonald U, Olson R, Beanan J, Russo TA. Metabolite Transporter PEG344 Is Required for Full Virulence of Hypervirulent Klebsiella pneumoniae Strain hvKP1 after Pulmonary but Not Subcutaneous Challenge. Infect Immun. 2017;85(10):e00093–17. Published 2017 Sep 20. doi:10.1128/IAI.00093-17

37. Yu F, Lv J, Niu S, Du H, Tang YW, Pitout JDD, et al. Multiplex PCR Analysis for Rapid Detection of Klebsiella pneumoniae Carbapenem-Resistant (Sequence Type 258 [ST258] and ST11) and Hypervirulent (ST23, ST65, ST86, and ST375) Strains. J Clin Microbiol. 2018 Aug 27;56(9):e00731–18.

38. Compain F, Babosan A, Brisse S, Genel N, Audo J, Ailloud F, et al. Multiplex PCR for detection of seven virulence factors and K1/K2 capsular serotypes of *Klebsiella pneumoniae*. J Clin Microbiol. 2014;52(12):4377–83.

39. Cai Y, Wang W, Liang H, Huang Q, Qin J, Guo Z, et al. Sensitive and specific LAMP and multiplex qRT-PCR assays for detection of hypervirulent Klebsiella pneumoniae. J Microbiol Methods. 2022;198:106480.

