## Supplementary Figure a-f, Agarose gel electrophoresis images showing validation and sensitivity of the m-PCR for "Multiplex PCR assay for the Rapid detection of *Klebsiella pneumoniae* pathotypes"

### Supplementary Figures

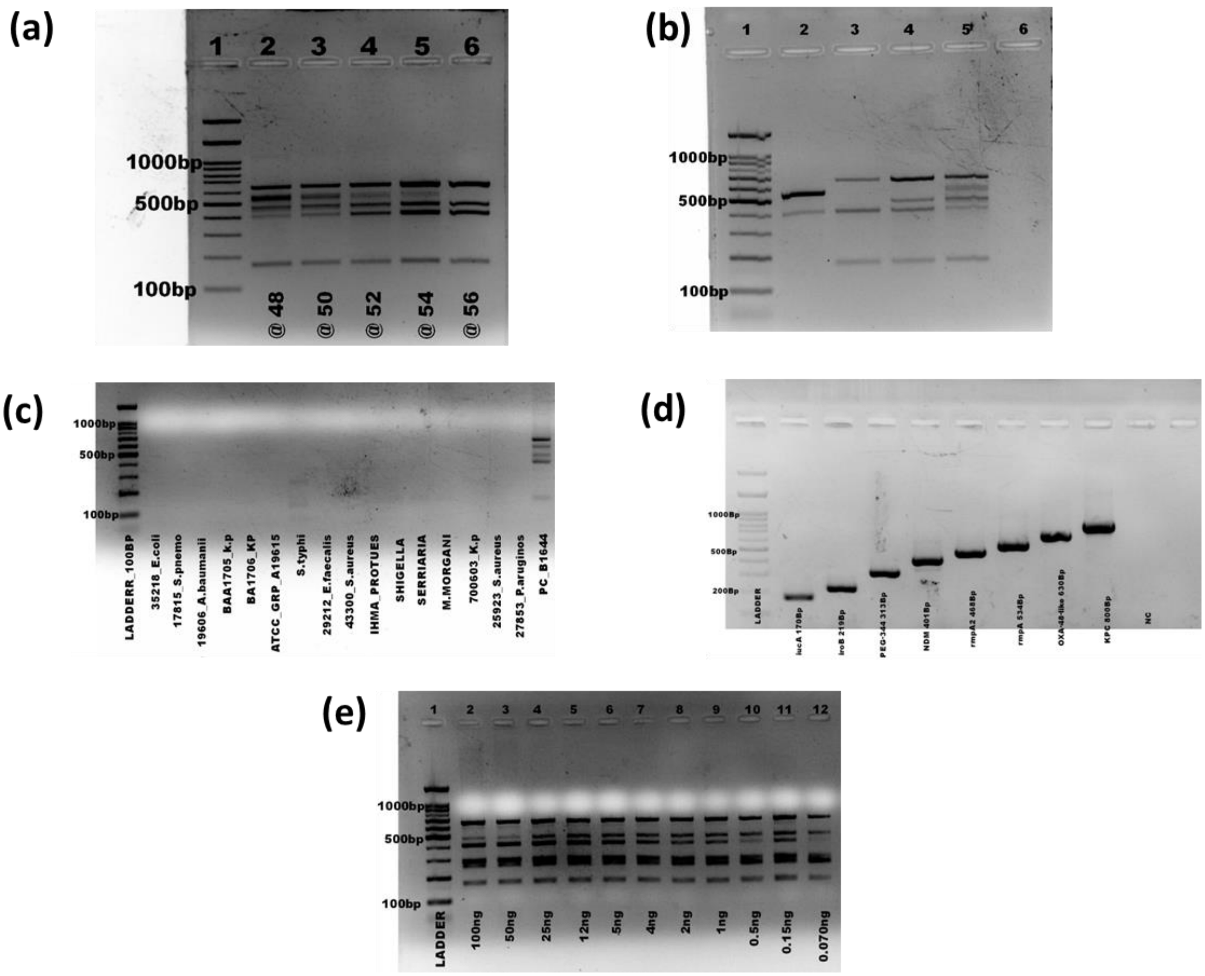


**Supplementary Figure a.** Determination of annealing temperature of the multiplex PCR. Lane 1: 1000 bp DNA marker; Lane 1: 48°C annealing; Lane 2: 50°C annealing; Lane 3: 52°C annealing; Lane 4: 54°C annealing; Lane 5:56°C annealing. **Figure b.** Optimization of the primers of m-PCR. Lane 1: 1000 bp DNA marker; Lane 1: duplex PCR; Lane 2: triplex PCR; Lane 3: quadruplex PCR; Lane 4: pentaplex PCR formed by *iucA, bla*$_{NDM}$, *rmpA, rmpA2* and *bla*$_{OXA\text{-}4\text{-}like}$. **Figure c**. Determination of specificity of the multiplex PCR. Lane 1: 1000 bp DNA marker, Lane 2-16: the template 14 bacterial strains of m-PCR which are negative. Lane 17: positive control. **Figure d.** Amplification of target gene of the multiplex PCR. Lane 1: 1000 bp DNA marker; Lanes 2–9: the template of single genes amplification respectively for *iucA, iroB, peg-344, bla*$_{NDM}$, *rmpA2, rmpA*, *bla*$_{OXA\text{-}48\text{-}like}$ *and bla*$_{KPC.}$ **Figure e**. Determination of the sensitivity of the multiplex PCR for bacterial genomic DNA detection for strain B1644, Lane 1: 1000 bp DNA marker; a Lanes 2–12: the concentration of Klebsiella pneumoniae DNA were 100 ng, 50 ng, 25 ng, 12 ng, 5 ng, 4 ng, 1 ng, 500 pg, 150 pg, and 70 pg respectively.

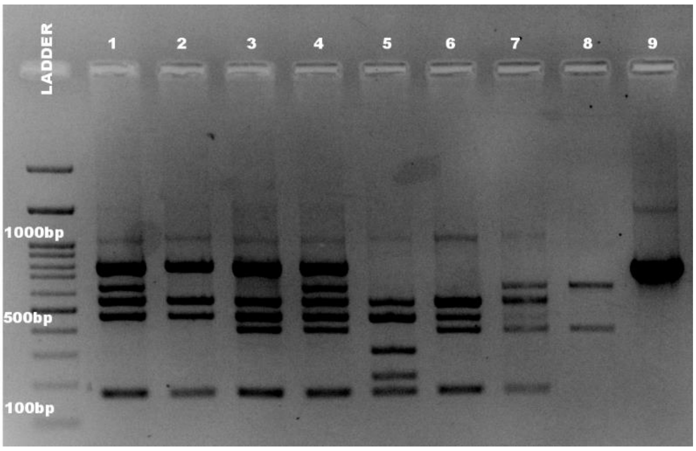


***Figure f.*** We evaluated various multiplex PCR combinations to determine the optimal setup for detecting key virulence (*rmpA, rmpA2, iucA, iroB, peg-344*) and resistance ($bla_{NDM}$, $bla_{OXA\text{-}48\text{-}like}$, *blaKPC*) genes. Different gene combinations were tested to assess their compatibility within a single assay. Lane-wise, the results are as follows: the first lane contains the molecular size marker (ladder), followed by multiplex amplification of *rmpA, rmpA2, iucA*, $bla_{OXA\text{-}48\text{-}like}$, $bla_{KPC}$ (Lane 1), *rmpA, rmpA2, iucA*, $bla_{KPC}$, $bla_{NDM}$ (Lane 2), and *rmpA, rmpA2, iucA*, $bla_{KPC}$, $bla_{NDM}$, $bla_{OXA\text{-}48\text{-}like}$ (Lane 3). Lane 4 includes the extended panel with *rmpA, rmpA2, iucA, iroB, and peg-344*, while Lane 5 and Lane 6 represent multiplex combinations with *rmpA, rmpA2, iucA*, $bla_{NDM}$ and *rmpA, rmpA2, iucA*, $bla_{NDM}$, $bla_{OXA\text{-}48\text{-}like}$, respectively. Lane 7 shows amplification of $bla_{OXA\text{-}48\text{-}like}$ and $bla_{NDM}$ alone, whereas Lane 8 confirms the presence of *blaKPC* individually. These results highlight the feasibility of a robust multiplex PCR approach for detecting hypervirulence and carbapenem resistance markers in *Klebsiella pneumoniae*, with additional genes available for extended analysis as needed.
